# Beta infection combined with Pfizer BNT162b2 vaccination leads to broadened neutralizing immunity against Omicron

**DOI:** 10.1101/2022.04.15.22273711

**Authors:** Sandile Cele, Mallory Bernstein, Farina Karim, Khadija Khan, Yashica Ganga, Zesuliwe Jule, Kajal Reedoy, Gila Lustig, Natasha Samsunder, Matilda Mazibuko, Shi-Hsia Hwa, Nithendra Manickchund, Anne von Gottberg, Nombulelo Magula, Richard J. Lessells, Tulio de Oliveira, Salim S. Abdool Karim, Willem Hanekom, COMMIT-KZN Team, Bernadett I. Gosnell, Mahomed-Yunus S. Moosa, Alex Sigal

## Abstract

Omicron (B.1.1.529) shows extensive escape from vaccine immunity, although vaccination reduces severe disease and death^1^. Boosting with vaccines incorporating variant spike sequences could possibly broaden immunity^2^. One approach to choose the variant may be to measure immunity elicited by vaccination combined with variant infection. Here we investigated Omicron neutralization in people infected with the Beta (B.1.351) variant and subsequently vaccinated with Pfizer BNT162b2. We observed that Beta infection alone elicited poor Omicron cross-neutralization, similar to what we previously found^3^ with BNT162b2 vaccination alone or in combination with ancestral or Delta virus infection. In contrast, Beta infection combined with BNT162b2 vaccination elicited neutralization with substantially lower Omicron escape.

## Results and Discussion

South Africa had a Beta dominated infection wave November 2020 to May 2021^4^. We enrolled 18 Beta infected participants based on infection date who were later vaccinated with Pfizer BNT162b2 (Table S1). A pre-vaccine sample was taken a median of 29 days post-symptom onset, and a post-vaccine sample 31 days post-vaccination. Five participants received one vaccine dose and the others two doses. None were boosted.

We measured plasma neutralization capacity as FRNT_50_ using live virus neutralization. Post-vaccination, Beta virus neutralization increased 21.3-fold (95% CI 9.7-46.8, Fig 1a) from geometric mean titer (GMT) FRNT_50_ of 112 to 2385. Omicron/BA.1 FRNT_50_ pre-vaccination was 6, rising post-vaccination 69.3-fold (95% CI 32.0-150, Fig 1b) to 447. Pre-vaccination, Omicron/BA.1 neutralization was 17.3-fold lower than Beta virus neutralization (95% CI 11.2-26.1, Fig 1c), confirming previous results^5^. Because many pre-vaccination samples did not reach 50% BA.1 neutralization with the most concentrated plasma tested, we tested additional samples from Beta infected individuals and obtained similar results (19.2-fold escape, Fig S1). In contrast, the fold-drop between Beta and Omicron/BA.1 neutralization post-vaccination was 5.3-fold (95% CI 4.4-6.4, Fig 1d). Hence, Omicron/BA.1 escape was reduced more than 3-fold. Similarly to BA.1, cross-neutralization of Omicron/BA.2 post-vaccination was only 4.2-fold lower (95% CI 3.4-5.2, Fig 1e). Ancestral virus neutralization showed a 1.5-fold decline relative to Beta (95% CI 1.2-1.8, Fig 1f).

**Figure 1:**
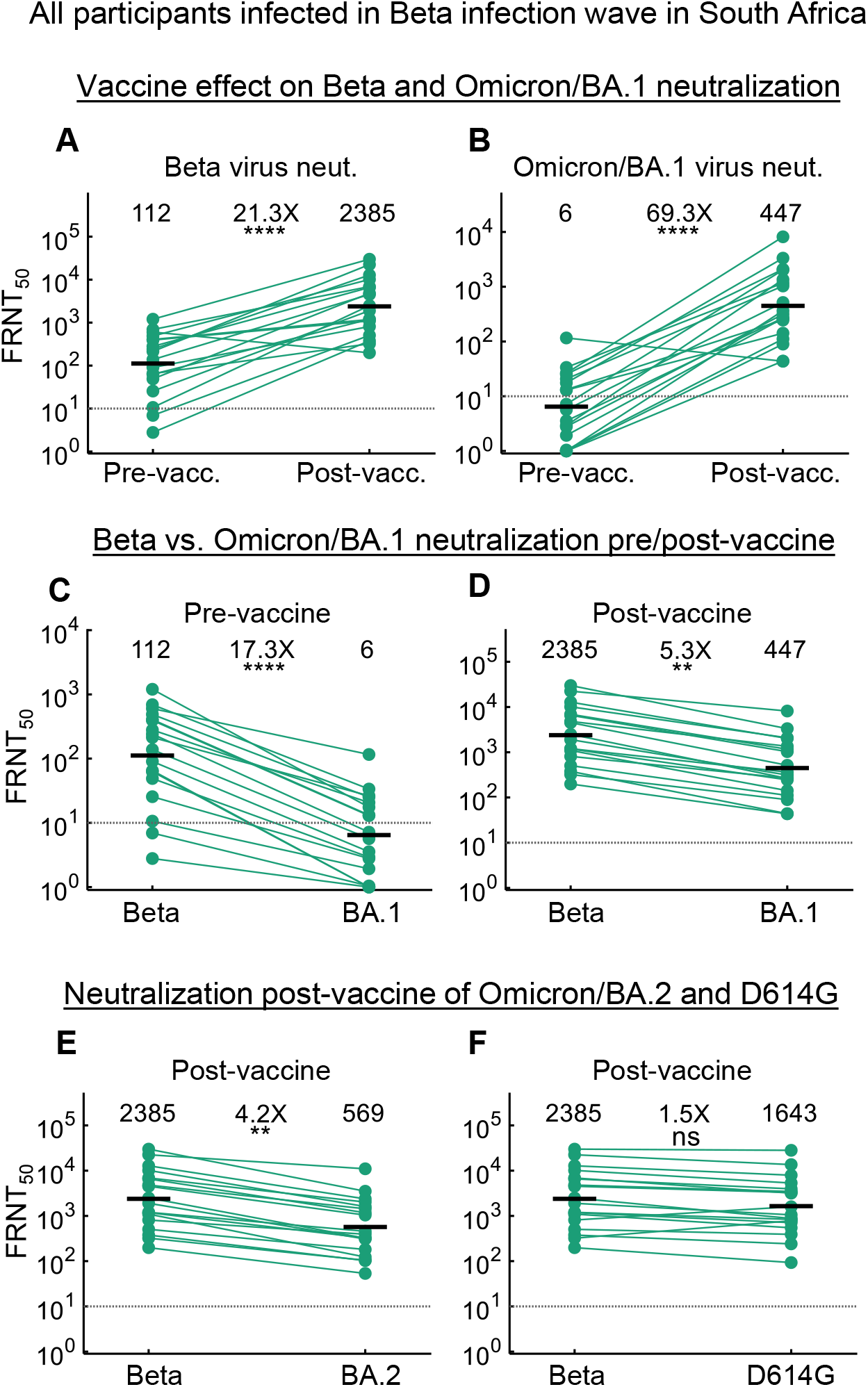
Hybrid immunity elicited by Beta variant infection combined with Pfizer BNT162b2 vaccination reduces Omicron escape. (A) Neutralization of Beta virus by plasma from n=18 convalescent participants infected during the Beta infection wave (December 2020 - May 2021) in South Africa, before (left) and after vaccination with BNT162b2 (right). Participants were sampled a median of 29 days post-symptom onset (pre-vaccine), and 31 days post-vaccine (Table S1). The same participant samples are used in the subsequent panels. Numbers are geometric mean titers (GMT) of the reciprocal plasma dilution (FRNT_50_) resulting in 50% neutralization. Fold-change is calculated by dividing the larger GMT value by the smaller value. Dashed line is most concentrated plasma tested. (B) Neutralization of Omicron/BA.1 virus before and after vaccination. (C) Neutralization of Beta versus Omicron/BA.1 virus pre-vaccination. (D) Neutralization of Beta versus Omicron/BA.1 virus post-vaccination. (E) Neutralization of Beta versus Omicron/BA.2 virus post-vaccination. (F) Neutralization of Beta versus ancestral/D614G virus post-vaccination. Data points are geometric means of FRNT_50_ measurements from two (BA.1 pre- and post-vaccine, Beta pre-vaccine, and BA.2 post-vaccine conditions) or four (Beta and D614G post-vaccine conditions) independent experiments performed on different days. p-values were: (A) 1.2 *×*10^*−*5^, (B) 3.1 *×*10^*−*7^, (C) 4.1 *×*10^*−*5^, (D) 0.0021, (E) 0.0050, (F) 0.10 as determined by the Wilcoxon rank sum test.

These results contrast with our previous findings for BNT162b2 vaccination alone or in combination with ancestral or Delta virus infection, where we observed approximately 20-fold drops in Omicron neutralization^3^. The limitations are that here we tested immunity generated by Beta virus infection followed by vaccination. Whether boosting with Beta sequence post-primary vaccination would be similar is unclear. However, it may be one line of evidence which argues for testing this approach.

## Data Availability

All data produced in the present study are available upon reasonable request to the authors.

## Supplementary appendix to

**Table S1:**
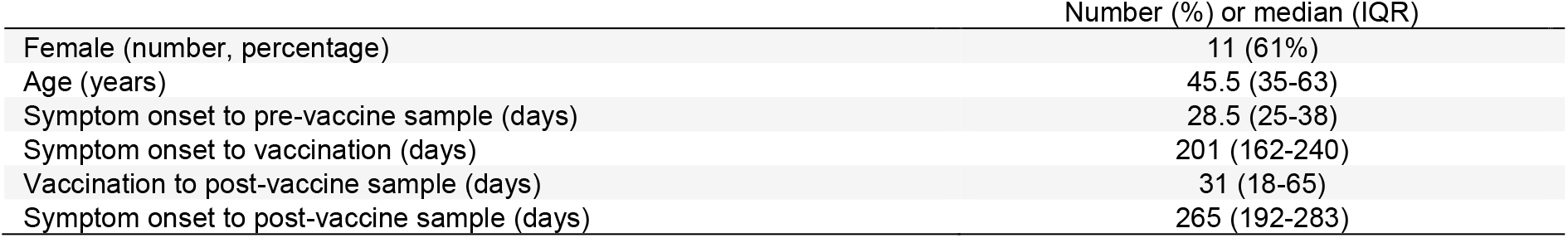
Summary participant characteristics

|  | Number (%) or median (IQR) |
| --- | --- |
| Female (number, percentage) | 11 (61%) |
| Age (years) | 45.5 (35-63) |
| Symptom onset to pre-vaccine sample (days) | 28.5 (25-38) |
| Symptom onset to vaccination (days) | 201 (162-240) |
| Vaccination to post-vaccine sample (days) | 31 (18-65) |
| Symptom onset to post-vaccine sample (days) | 265 (192-283) |

### Supplementary materials and methods

#### Informed consent and ethical statement

Blood samples used for plasma isolation and neutralization experiments and swabs for isolation of the ancestral/D614G and Delta viruses were obtained after written informed consent from adults with PCR-confirmed SARS-CoV-2 infection who were enrolled in a prospective cohort study at the Africa Health Research Institute approved by the Biomedical Research Ethics Committee at the University of KwaZulu–Natal (reference BREC/00001275/2020). The Beta virus was obtained from residual swab samples used for diagnostic testing by the National Health Laboratory Service (BREC approval reference BREC/00001510/2020). The Omicron/BA.1 virus was isolated from a residual swab sample with SARS-CoV-2 isolation from the sample approved by the University of the Witwatersrand Human Research Ethics Committee (HREC) (ref. M210752). The sample to isolate Omicron/BA.2 was collected after written informed consent as part of the “COVID-19 transmission and natural history in KwaZulu-Natal, South Africa: Epidemiological Investigation to Guide Prevention and Clinical Care” Centre for the AIDS Programme of Research in South Africa (CAPRISA) study and approved by the Biomedical Research Ethics Committee at the University of KwaZulu–Natal (reference BREC/00001195/2020, BREC/00003106/2021).

#### Data and sequence availability statement

Sequence of Omicron sub-lineage viruses have been deposited in GISAID with accession EPI_ISL_7886688 (Omicron/BA.1), EPI_ISL_9082893 (Omicron/BA.2), EPI_ISL_678615 (Beta), and EPI_ISL_602622 (ancestral D614G). Raw images of the data are available upon reasonable request.

#### Cells

H1299 cell lines were propagated in growth medium consisting of complete Roswell Park Memorial Institute (RPMI) 1640 medium with 10% fetal bovine serum containing 10mM of HEPES, 1mM sodium pyruvate, 2mM L-glutamine and 0.1mM nonessential amino acids. H1299 cells were passaged every second day. The H1299-E3 (H1299-ACE2, clone E3) cell line was derived from H1299 (CRL-5803) as described in our previous work^1,2^.

#### Live virus neutralization assay

H1299-E3 cells were plated in a 96-well plate (Corning) at 30,000 cells per well 1 day pre-infection. Plasma was separated from EDTA-anticoagulated blood by centrifugation at 500 rcf for 10 min and stored at −80 °C. Aliquots of plasma samples were heat-inactivated at 56 °C for 30 min and clarified by centrifugation at 10,000 rcf for 5 min. Virus stocks were used at approximately 50-100 focus-forming units per microwell and added to diluted plasma.

Antibody–virus mixtures were incubated for 1 h at 37 °C, 5% CO_2_. Cells were infected with 100 μL of the virus–antibody mixtures for 1 h, then 100 μL of a 1X RPMI 1640 (Sigma-Aldrich, R6504), 1.5% carboxymethylcellulose (Sigma-Aldrich, C4888) overlay was added without removing the inoculum. Cells were fixed 18 h post-infection using 4% PFA (Sigma-Aldrich) for 20 min. Foci were stained with a rabbit anti-spike monoclonal antibody (BS-R2B12, GenScript A02058) at 0.5 μg/mL in a permeabilization buffer containing 0.1% saponin (Sigma-Aldrich), 0.1% BSA (Sigma-Aldrich) and 0.05% Tween-20 (Sigma-Aldrich) in PBS. Plates were incubated with primary antibody overnight at 4 °C, then washed with wash buffer containing 0.05% Tween-20 in PBS. Secondary goat anti-rabbit HRP conjugated antibody (Abcam ab205718) was added at 1 μg/mL and incubated for 2 h at room temperature with shaking. TrueBlue peroxidase substrate (SeraCare 5510-0030) was then added at 50 μL per well and incubated for 20 min at room temperature. Plates were imaged in an ImmunoSpot Ultra-V S6-02-6140 Analyzer ELISPOT instrument with BioSpot Professional built-in image analysis (C.T.L).

### Statistics and fitting

All statistics and fitting were performed using custom code in MATLAB v.2019b. Neutralization data were fit to:

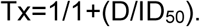

Here Tx is the number of foci normalized to the number of foci in the absence of plasma on the same plate at dilution D and ID_50_ is the plasma dilution giving 50% neutralization. FRNT_50_ = 1/ID_50_. Values of FRNT_50_ <1 are set to 1 (undiluted), the lowest measurable value. We note that the most concentrated plasma dilution was 1:10 for experiments with weak neutralization (pre-vaccine samples) and therefore FRNT_50_ < 10 were extrapolated. To calculate confidence intervals, FRNT_50_ or fold-change in FRNT_50_ per participant was log transformed and arithmetic mean + 2 std and arithmetic mean - 2 std were calculated for the log transformed values. These were exponentiated to obtain the upper and lower 95% confidence intervals on the geometric mean FRNT_50_ or the fold-change in FRNT_50_ geometric means.

## Acknowledgements

This study was supported by the Bill and Melinda Gates award INV-018944 (AS), National Institutes of Health award R01 AI138546 (AS), and South African Medical Research Council award 6084COAP2020 (AS). The funders had no role in study design, data collection and analysis, decision to publish, or preparation of the manuscript.

---

**Figure S1:**
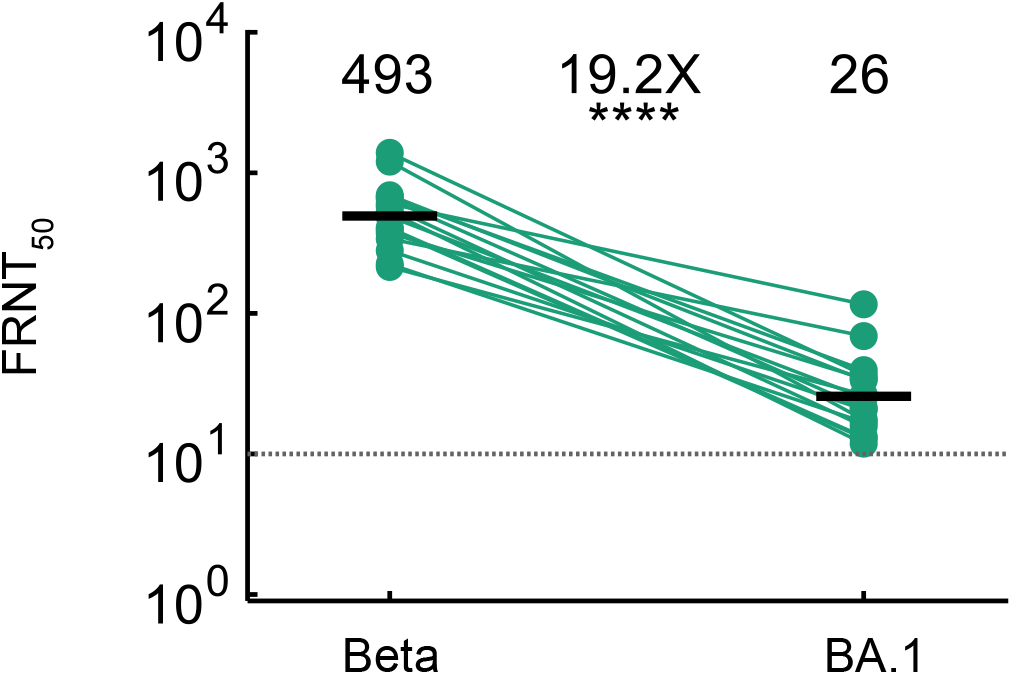
Omicron escape in participants with Omicron neutralization greater than FRNT_50_=10. For this analysis, participant samples pre-vaccination (n=8, participant numbers 1, 5, 6, 12, 13, 14, 17, 18 according to Table S2) were selected based at least 50% Omicron/BA.1 neutralization at the most concentrated plasma used (1:10 dilution, corresponding to FRNT_50_=10 shown by dashed horizontal line). In addition, n=8 samples from unvaccinated participants infected in the Beta infection wave in South Africa were selected solely on FRNT_50_>10 for Omicron/BA.1 neutralization (participants 19-26 listed on Table S3). Data points are geometric means of FRNT_50_ measurements from two independent experiments performed on different days. p-value was 7.7×10^−7^ as determined by the Wilcoxon rank sum test and fold-change 95% confidence intervals were 14.1 to 28.1-fold.

